# Causally-informative analyses of the effect of job displacement on all-cause and specific-cause mortality from the 1990s Finnish recession until 2020: A population registry study

**DOI:** 10.1101/2024.08.30.24312834

**Authors:** Stephanie Zellers, Elissar Azzi, Antti Latvala, Jaakko Kaprio, Terhi Maczulskij

**Affiliations:** Institute for Molecular Medicine Finland, University of Helsinki, Helsinki, Finland; Institute of Criminology and Legal Policy, University of Helsinki, Helsinki, Finland; ETLA Economic Research, Helsinki, Finland; IZA Institute of Labor Economics, Bonn, Germany

## Abstract

**Background:** Job loss is related to morbidity and mortality, but generation of causal evidence is challenging due to confounding factors. Finland suffered a severe economic recession in the early 1990s with unemployment reaching almost 25%, with many job losses due to mass layoff or company closure. Such job displacements are assumed to be exogenous to the individual and create a natural experiment for causal inference.

**Methods:** We evaluate the causal relationship between job displacement and mortality using register data from Finland between 1988-2020 (N=590,823 individuals [43·3 % female] aged 25-55 and securely employed by the private sector at baseline, N=93,199 total deaths by 2020).

**Findings:** Job displacement is associated with increased risk of all-cause mortality even after accounting for sex, age, marital status, and education (HR=1·09 [1·07, 1·10]). Risks of death by suicide, violence, alcohol, accidents, and disease are higher for displaced individuals at all follow-up periods examined. Risks of death from cancer and ischaemic heart disease are higher for displaced individuals only in later follow up periods.

**Interpretation:** Our analyses support the causal influence of job displacement on all-cause and specific-cause mortality, even up to 30 years after the recession; this risk varies by cause of death and by length of follow-up. Future work should evaluate stress and substance use as potential pathways from job displacement to mortality.

**Funding:** This work was supported by the Biology of Trauma Initiative at the Broad Institute of MIT and Harvard and the Academy of Finland Centre of Excellence in Complex Disease Genetics.

**Research in Context:** *Evidence before this study:* Job loss and unemployment are stressful life events. We searched PubMed from database inception until August 24, 2024 with the search terms “(mortality) AND (job loss) OR (job displacement)” to identify research utilizing natural experiment job displacement designs to estimate the effect of job loss on all-cause and/or cause-specific mortality. No restrictions were applied to the language of publication or article type. We also reviewed references to identify additional relevant studies. Meta-analytic estimates indicate that job loss negatively influences mental and physical health and confers risk for mortality. That said, unemployment and health have a complex relationship, where poor health can increase likelihood of job loss, which then in turn worsens health; therefore, natural experiments are required for causal inference. Causally-informative studies of job loss suggest that job loss causally increases risk of all-cause mortality up to 20 years after job loss and some specific causes of death within 5-10 years after job loss.

*Added value of this study:* This study expands the length of follow-up after job loss (up to 30 years) and evaluates a broader range of specific causes of death, such as violence. The long follow-up permits investigation into deaths resulting from diseases that develop over long periods of time, like heart disease and cancer.

*Implications of all available evidence:* Job loss causally increases risk of all-cause and specific-cause mortality and the risk of mortality from specific causes varies by duration after job loss, with external causes of death showing increased risk shortly after job loss and long-term illnesses like cancer or heart disease showing increased risk long after the job loss event. Substance use and stress are implicated as possible mechanisms for future research.

## Introduction

Finland suffered a dramatic recession in the early 1990s as a result of lax financial policy and collapse of trade with the Soviet Union^1^. During this time, Finland’s GDP shrank by 14% and unemployment rose to over 20%. While the economy recovered across the rest of the decade, the consequences to public health have persisted.

Macroeconomic studies of recessions and economic growth find a procyclical fluctuation in mortality; in other words, unemployment and mortality have a negative relationship where mortality rates decrease as unemployment rates increase^2^. Further work suggests that while increasing unemployment rates may indirectly benefit health, personal experiences of economic crisis have a detrimental effect on health^3,4^.

The loss of a job is one example of negative personal impacts during a recession. Job loss, even in the absence of an extended period of unemployment, is a stressful life event and is often followed by additional stressful events like financial strain, relocation, or change in social status^3,5,6^. Job loss and unemployment are associated with negative health effects, but the relationship between employment and health may be partly driven by health-related selection effects and there may be a two-way relationship between unemployment and health^3,4,7–9^. Advanced research designs are needed to disentangle the causal mechanisms involved.

Displacement designs are a quasi-experimental analysis used to draw causally-informative conclusions about the effects of job loss. In a displacement design, the exposure is involuntary job loss through plant closure or downsizing^10,11^. These designs rely on the assumption that job loss from plant closure or downsizing is exogenous to the individual who loses their job, and therefore the relationship between job loss and the outcome of interest is less confounded by endogenous factors like health or other personal characteristics. Furthermore, when administrative data are available, involuntary job losses can be identified in a systematic and objective manner and in recessions, involuntary displacements become more commonplace.

A variety of studies based on administrative data have evaluated the effect of job displacement on health and mortality in recession and non-recession contexts. Job displacement increases the risk of all-cause mortality by anywhere from 10-20%, up to 20 years after displacement and this effect has been established in Denmark^12^, the Netherlands^13^, Sweden^14^, Finland^15,16^, and the United States^17^. This effect is specific to displaced individuals; those who remained employed in workplaces that downsized are not at higher risk of mortality^16^.

Job displacement is also associated with increased risk of specific causes of death such as alcohol^12,14,15,18^, substance use^19^, cardiovascular/circulatory disease^12,15^, suicide^12,14,15^, traffic accidents^12^, and mental illness^12^ up to 20 years after displacement. We expand this field of research with a large sample, long follow-up duration, and broader range of specific causes of death.

Here we capitalize on administrative data before, during, and after the 1990s recession in Finland to create a large cohort of involuntarily displaced individuals and non-displaced controls with 30 years of follow-up. Using this natural experiment, we evaluate the influence of job loss during the 1990s Finnish economic recession on mortality until 2020. Our work expands on prior research through the long duration of follow-up, which is crucial for diseases that may take longer to develop, as well as thorough investigation of specific causes of death. Lastly, we are able to utilize the entire Finnish population, as opposed to earlier works that needed to use a subsample of all Finnish residents^16,18,19^.

Our research questions were as follows: 1) What is the relationship between job displacement during the 1990s Finnish economic recession and all-cause mortality until 2020? 2) What are the relationships between job displacement and specific causes of death? These analyses were pre-registered; the analysis plan (including any changes) is available at https://osf.io/bxydp/.

## Methods

### Sample

The study base consists of all individuals residing in Finland at some point between the years of 1988-1994, according to the Digital and Population Data Services Agency of Finland. These persons were linked using the unique personal identifier assigned to all citizens and residents to register information at Statistics Finland. This includes employment history, migration, date and cause of death, education, civil status, birthdate, and sex assigned at birth.

We then utilized linked employee-employer data to restrict the sample to full-year (12 months of employment), non-student wage earners between the ages of 25-55 who had been employed continuously for at least two years in private-sector companies with at least 10 employees. These inclusion criteria reduce endogeneity issues by identifying a sample of individuals of prime working age that were securely attached in the labor market prior to the recession’s start. We excluded public sector workers because their employer codes are not well defined by Statistics Finland. These criteria resulted in a final analytic sample of N=590,823 individuals.

### Exposure

The primary exposure is involuntary job loss (henceforth referred to as job displacement) during Finland’s 1990s severe economic recession. To minimize confounding factors, we operationalized job displacement as a binary variable indicating exogenous and involuntary job loss using linked employer-employee data. We define displaced workers as individuals separated from their private-sector jobs following plant closures or mass lay-offs of 50% or more employees. To address potential endogeneity issues due to firm restructuring, plant closures were not classified as genuine if a worker obtains a position within the same firm subsequently, or a significant proportion (70%) of displaced workers from the same plant relocate to another plant owned by the same firm within the next year^6^. This distinction differentiates between genuine plant closures and other organizational changes.

The job displacement exposure was coded for each year during the recession (observation years of 1990, 1991, 1992, 1993, 1994), resulting in N=1,549,081 total observations. The exposed sample is comprised of the individuals who experienced at least one job displacement during the recession (N=114,257). As the control group, we use the sample of workers meeting the same employment and age criteria who were not displaced at any point during the recession (N=476,566).

Although job displacements typically do not accumulate for the same individuals, it is observed that less-skilled workers tend to match with weaker firms and approximately 3% of displaced individuals in the sample experienced more than one displacement^10^. To account for this potential bias, we focus only on the first displacement event for a given individual. This exposure definition has been validated in previous economic papers^6,10^.

### Outcomes

Our primary outcome was all-cause mortality. We defined a death as individuals who died by the last calendar day in the target follow-up period (ex. December 31, 2020).We defined several cumulative follow up periods to align with follow-up durations in previous literature: 1990-2000, 1990-2005, 1990-2010, 1990-2020.

Our second outcome of interest was the underlying cause of death. The Supplemental Methods and Table S1 details the classifications of causes of death utilized in the Finnish Death Register. Causes of particular interest, given their relation to life stressors, were suicide, cancer, ischaemic heart diseases, diseases and accidental poisoning by alcohol, violence, and accidents. We also included causes that we did not expect to be related to job displacement, such as dementia, infectious and respiratory diseases, and endocrine disorders as negative controls.

### Covariates

We utilized covariates from the register data, selected either due to their relationship to mortality, labor market outcomes, or both. Covariates were coded for the year prior to the observation year. We covaried for sex (male or female, as assigned at birth), education, marital status (married or not), and whether the individual had children under the age of 18 (yes or no). Education was classified as primary, secondary or tertiary (see Supplemental Methods for details). Statistics Finland does not produce statistics on race or ethnicity, only nationality and native language, therefore we cannot produce descriptives on race. Instead we describe native language and background. Finnish residents, born in Finland or abroad, are considered to have “Finnish background” based on having one of the three Finnish national languages (Finnish, Swedish, or Sami) as their native language^20^.

### Analyses

#### Descriptives

We first described the data by generating frequencies, means, standard deviations, and/or ranges for our exposure, outcomes, and covariates. We generated descriptives separated by displacement and control status as well as year.

#### All-Cause Mortality

To evaluate the relationship between job displacement and mortality, we computed Cox proportional hazards models using the R package “Survival”^21,22^. Death from all causes was predicted from job displacement and covariates, with age in years since birth as the underlying time metric. Persons aged 25 to 55 on January 1^st^, 1990, entered the analysis on January 1^st^, 1990, while those who turned 25 years of age between January 1^st^, 1990 and December 31^st^, 1994 entered on their 25^th^ birthday. Persons entering the analysis frame contributing non-exposed person-time until the end-of-follow-up (death or December 31^st^, 2020) unless they experienced job-displacement between January 1^st^, 1990 and December 31^st^, 1994. In that case they contributed exposed person-time from there onwards. Individuals were also censored at emigration, if there was no subsequent return to Finland nor post-emigration register-based information on their mortality status (N=4,840 censorings due to emigration).

We evaluated these survival models with varying durations of follow-up (1990-2000, 1990-2005, 1990-2010, 1990-2020) in order to evaluate how any associations may change over time. Models were run at the level of all observations (N=1,549,081 total observations) and were clustered by individuals to account for multiple observation years per person.

#### Cause-Specific Mortality

Death from a specific cause during the follow-up period was predicted as for overall mortality but with the specific cause of death as the outcome. Those not deceased contribute person-time at risk until the end of the follow-up period. Those deceased from other causes also contribute person-time at risk until their time of death from another cause^23^. This method treats competing risks as censoring events.

#### Supplemental Analyses

We evaluated various non-preregistered supplemental analyses (propensity score matching and stratified models) to further disentangle the relationships between displacement, mortality, and covariates and evaluate the robustness of our findings. Full descriptions of these analyses are presented in the Supplemental Methods.

## Results

### Descriptives

The analytic sample of N=590,823 individuals was 43·3% female and 99% of individuals were born in Finland; 93·7% of the sample were native Finnish speakers, 5·7% were native Swedish speakers, and 0·6% had a native language other than Finnish or Swedish. Of the N=5781 individuals born outside of Finland, 31·6% were individuals of Finnish background and 74·2% had Finnish nationality by the time of entry in the study.

A total of N=114,257 individuals were displaced at least once during the recession representing 19·3% of the total sample and the frequency of displacements varied by year across the recession (See Table 1). Of the 114,257 individuals displaced at least once, 39·3% were women. Table 1 contains additional descriptives about each year of assessment and covariates for both the displaced and control samples.

**Table 1:** Descriptives of Sample Across Possible Years of Exposure.

|  | **1990** |  | **1991** |  | **1992** |  | **1993** |  | **1994** |  |
| --- | --- | --- | --- | --- | --- | --- | --- | --- | --- | --- |
|  | Controls | Displaced | Controls | Displaced | Controls | Displaced | Controls | Displaced | Controls | Displaced |
| N<br>Individuals | 282,929 | 15,071 | 294,592 | 31,194 | 297,858 | 31,569 | 284,045 | 22,132 | 275,400 | 14,291 |
| %<br>Female | 43.9 | 43.3 | 44.3 | 37.8 | 44.8 | 37.0 | 44.3 | 39.3 | 43.4 | 43.5 |
| Age Mean<br>(SD) | 40.0<br>(8.1) | 39.2<br>(8.4) | 40.0<br>(8.2) | 39.3<br>(8.3) | 40.1<br>(8.2) | 39.4<br>(8.3) | 40.2<br>(8.2) | 40.0<br>(8.3) | 40.5<br>(8.2) | 40.4<br>(8.2) |
| % Secondary<br>Education | 37.6 | 41.9 | 37.9 | 42.8 | 38.3 | 40.4 | 39.2 | 38.9 | 40.0 | 37.1 |
| % Tertiary<br>Education | 24.8 | 19.5 | 26.4 | 20.2 | 27.5 | 25.6 | 28.3 | 28.9 | 29.0 | 29.8 |
| % Married | 65.9 | 62.1 | 64.6 | 60.5 | 63.4 | 61.5 | 62.6 | 61.4 | 62.1 | 61.2 |
| % Children<br>Under 18 | 51.5 | 49.8 | 50.8 | 49.2 | 50.4 | 51.2 | 50.5 | 50.9 | 50.3 | 51.0 |

Note: Covariates are measured the year prior to exposure

There were N=93,199 deaths by December 31^st^, 2020, representing 15·8% of the sample. Median age at death was 65 years (SD=10·9 years, Range=26-85). The mean age of death in displaced individuals was 63·4, and the mean age of death in controls was 64·3, (95% CI age difference [0·72, 1·07]).Total deaths by end of follow-up from each specific cause are presented in Table S1. Cancer was the most common cause of death, whereas violence was the least common.

### All-Cause Mortality

Cox proportional hazards models revealed an increased risk of all-cause mortality in displaced individuals as compared to controls; this risk was elevated in all follow-up lengths but the effect size attenuated over time (adjusted model estimates: 1990-2000 HR=1·17; 1990-2005 HR=1·14; 1990-2010 HR=1·12; 1990-2020 HR=1·09). In addition to the effect of displacement, female sex, being married and having tertiary education were significantly associated with lower risk of mortality at all follow-up lengths. Results of the covariate-adjusted models are visualized in Figures 1 and 2, as well as Table 2; unadjusted model results are presented in Table 2.

**Figure 1.**
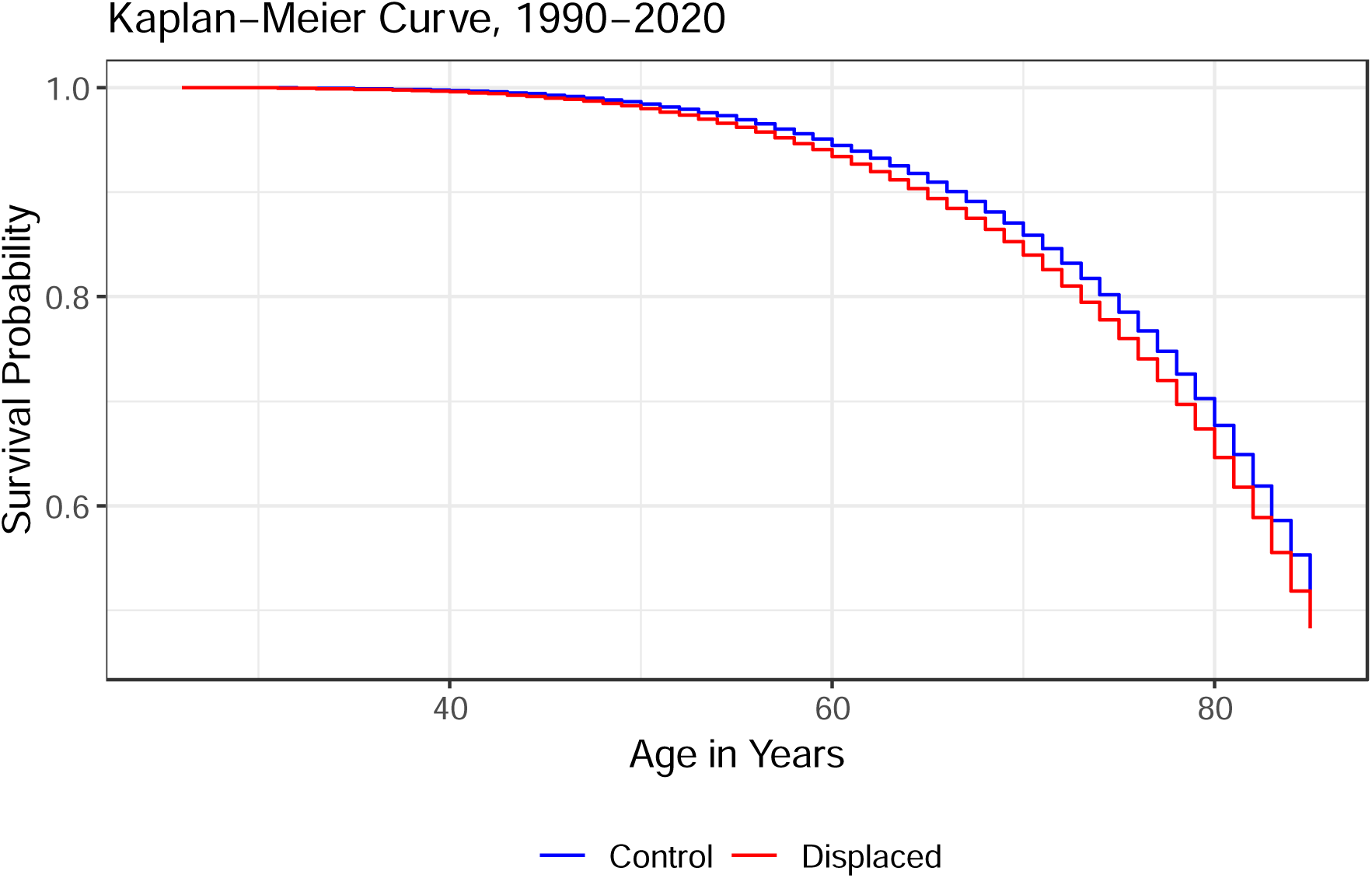
Kaplan-Meier Survival Curve for all-cause mortality by job displacement status, from 1990-2020.

**Figure 2.**
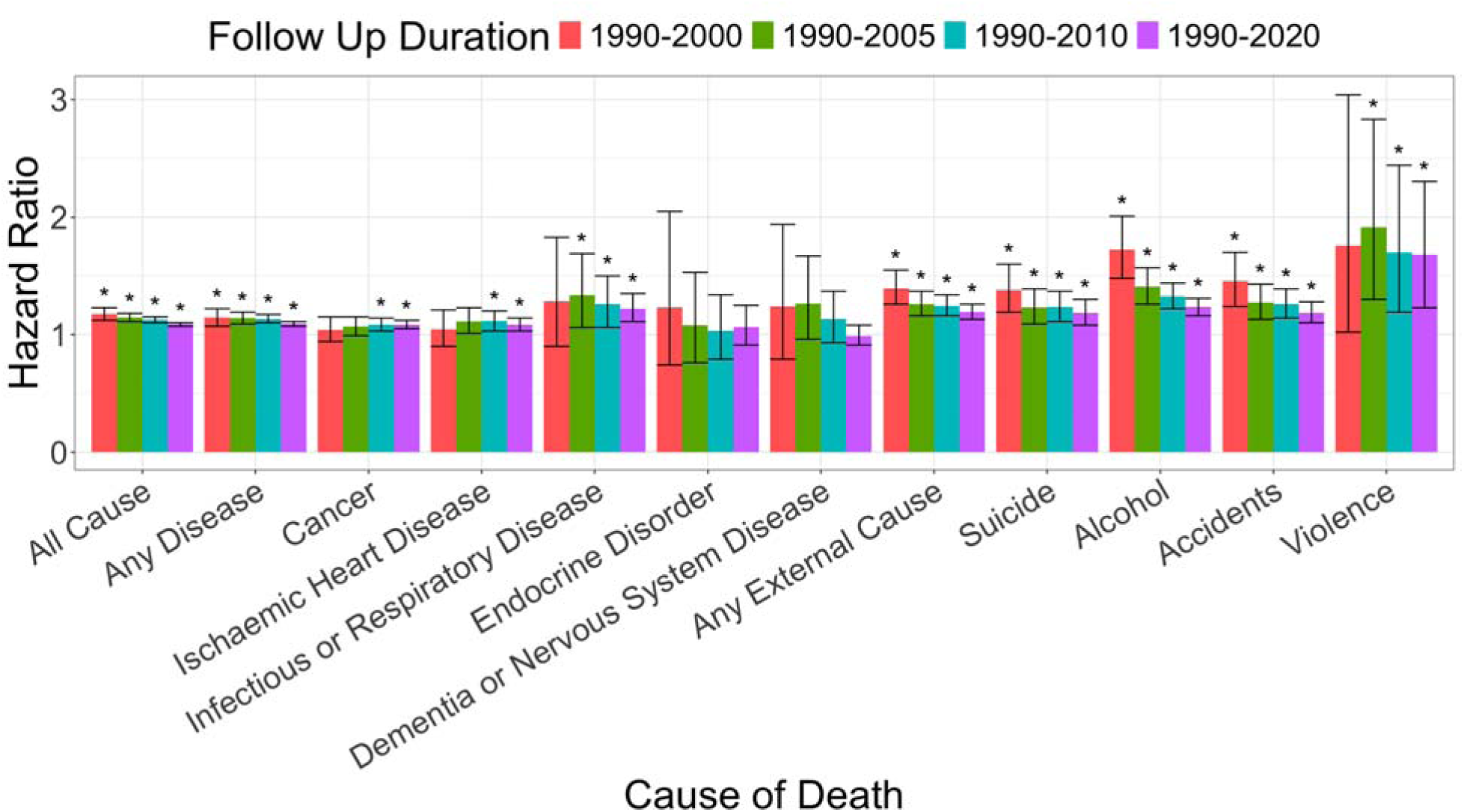
Hazard ratios representing the excess mortality risk after job displacement, organized by cause of death and follow-up duration. Bars represent 95% confidence intervals and * indicates HR that are significantly larger than 1.

**Table 2:** Results from All-Cause Mortality Survival Models.

| HR [95% CI] | Follow Up Duration |  |  |  |
| --- | --- | --- | --- | --- |
|  | 1990-2000 | 1990-2005 | 1990-2010 | 1990-2020 |
| **Unadjusted Model Estimates** |  |  |  |  |
| Displacement | 1.26<br>[1.20, 1.31] | 1.22<br>[1.18, 1.26] | 1.20<br>[1.17, 1.23] | 1.15<br>[1.13, 1.16] |
| **Adjusted Model Estimates** |  |  |  |  |
| Displacement | 1.17<br>[1.12, 1.23] | 1.14<br>[1.11, 1.18] | 1.12<br>[1.10, 1.15] | 1.09<br>[1.07, 1.10] |
| Sex | 0.41<br>[0.39, 0.43] | 0.42<br>[0.40, 0.43] | 0.43<br>[0.42, 0.45] | 0.48<br>[0.47, 0.49] |
| Secondary Education | 1.07<br>[1.02, 1.13] | 1.02<br>[0.99, 1.06] | 0.96<br>[0.94, 0.98] | 0.90<br>[0.89, 0.92] |
| Tertiary Education | 0.72<br>[0.68, 0.77] | 0.71<br>[0.68, 0.74] | 0.68<br>[0.65, 0.70] | 0.66<br>[0.64, 0.67] |
| Marital Status | 0.51<br>[0.49, 0.53] | 0.55<br>[0.53, 0.57] | 0.59<br>[0.57, 0.60] | 0.67<br>[0.66, 0.68] |
| Children Under 18 | 1.10<br>[1.05, 1.15] | 1.02<br>[0.99, 1.05] | 0.96<br>[0.94, 0.98] | 0.90<br>[0.89, 0.92] |

Note: The reference category for displacement was non-displaced individuals, the reference category for sex was male, the reference category for education was primary, the reference category for marital status was unmarried, and the reference category for children under 18 was no.

### Cause-Specific Mortality

We identified patterns of elevated risk of mortality from job displacement, which varied by cause and follow-up length (Figure 2). Full model estimates are presented in Table S2. The risks for mortality from accidents, alcohol, violence, and suicide were elevated in displaced individuals as compared to their non-displaced peers at all follow-up lengths.

Cancer and ischaemic heart disease only showed elevated risk in displaced peers at longer follow-up lengths (1990-2010 and 1990-2020). As expected, risks of death from endocrine and dementia/nervous system disorders were not significantly related to job displacement. Unexpectedly, displaced individuals were also at higher risk of death from infectious and respiratory disease at later follow-up lengths (1990-2005, 1990-2010, 1990-2020).

We investigated this finding further in a non-preregistered follow-up by disaggregating this broader cause and looking only at deaths due COPD, for which smoking is a major cause^24^. COPD represented 51% of deaths within the infectious and respiratory disease category. We correspondingly also isolated “lung” cancer deaths (including larynx, trachea, bronchus, and lung) for which smoking is the main cause^24^; these represented 21% of the total cancer deaths. Displacement significantly predicted mortality from COPD and lung cancer during the follow up periods 1990-2010 (lung cancer HR=1·14 [1·01, 1·28]; COPD HR=1·37 [1·02, 1·83]) and 1990-2020 lung cancer HR=1·21 [1·14, 1·30]; COPD HR=1·21 [1·05, 1·38]), indicating the need for mechanistic follow-up work on smoking and related lifestyle factors after displacement.

### Supplemental Analyses

A balance check for the propensity score matching is presented in Table S3; this table corresponds to Table 1 in the full sample. Results of the all-cause mortality survival models in the propensity score matched sample are presented in Table S4. Overall, the results from the propensity score analysis agree with respect to effect significance and direction as compared to the analysis in the full sample, though the effect sizes for job displacement are somewhat attenuated. This propensity-score matching therefore further supports the existence of a causal impact of job displacement on mortality.

A more detailed description of the stratified model results is presented in the Table S5. Stratifying by education did not produce different estimates as compared to the primary models including education as a covariate, even though education predicts mortality. Stratifying by sex indicated that the effect sizes for displacement are larger for males as compared to females, in addition to the significant effect of sex as a covariate in the primary model.

Birth years ranged from 1935-1969 and therefore age at displacement varied within the sample; effect sizes for displacement were smallest for individuals born 1935-1946 and largest for individuals born 1959-1969. Lastly, stratifying by year of displacement indicated that the effect of displacement varied greatly by year. Individuals displaced in 1994 showed no increased risk of mortality, whereas individuals displaced in 1990 or 1991 were at the highest risk of mortality. Individuals displaced in 1992 or 1993 had elevated risk of mortality, though not as large as those displaced in 1990 or 1991.

## Discussion

We evaluated the effect of involuntary exogenous job loss on all-cause and specific-cause mortality in Finnish private sector employees during the 1990s recession using administrative data. We extend previous research with a longer duration of follow up (1990-2020), larger sample size, and expanded list of specific causes of death.

We demonstrate comparable findings to previous studies indicating that risk of all-cause mortality is raised by approximately 10-20% and that the effect persists long after the displacement event^12–17^. Additionally, our specific causes of death findings align with existing literature showing increased risk of mortality due to alcohol^12,14,15,18^, ischaemic heart disease^12,15^, suicide ^12,14,15^, and accidents^12^. Our results also agree with previous research that suggests stress as a mechanism between job displacement and death^13^. That said, studies of hospitalizations for stress-related diseases of the circulatory and digestive systems have not been shown to significantly increase after job displacement in Danish administrative data up to five years after displacement^25^. The link between job displacement, stress, and morbidity/mortality requires further study; long follow-up durations may be necessary to observe effects in stress-related diseases that develop later in life.

We report novel results with respect to causes of death, specifically of interest are deaths due to all cancers and infectious/respiratory disease that have not been previously established. We could not locate studies of job displacement that investigated infectious and respiratory disease as a specific cause of death after displacement.

Studies on cancer are sparse. Browning and Heinesen (2012) did not find any significant increase in all-cancer mortality after job displacement in Danish administrative data even in up to 20 years of follow up^12^, which is in contrast with our finding of increased all-cancer mortality up to 20 and 30 years after displacement. However, when we analyzed lung cancer deaths and COPD, we find that displaced individuals are at higher risk of lung cancer and COPD mortality. This is consistent with prior work indicating that smoking increases in displaced individuals^26^. Despite not finding significantly increased all-cancer mortality risk, Browning and Heinesen (2012) also identified significantly increased risk of death from smoking related cancers^12^.

Lastly, the supplemental stratified models suggested differential effects of job displacement on all-cause mortality risk by demographic group. Effect sizes for all-cause mortality risk after displacement were larger in males as compared to females. In other words, males have higher risk of mortality as compared to females regardless of displacement, but the effect of displacement on mortality risk is also higher in males as compared to females. This is in addition to the main effect of sex on all-cause mortality risk and after accounting for age. Additionally, the effect of displacement on mortality varied by birth and displacement years, with displacements conferring more risk early in the recession as opposed to later and displacements conferring greater risk to younger individuals as compared to older. That said, we cannot disentangle age and cohort effects here. Conversely, we did not find evidence that the effect of displacement on all-cause mortality risk varied by education level, though education does predict all-cause mortality risk as a main effect.

In total, men are at higher risk of all-cause mortality as compared to females, men are more likely to experience job displacement as compared to females, and the effect of job displacement on all-cause mortality is stronger in males as compared to females. Timing of displacement, both the year of displacement and age of the displaced individual, also confers additional risk.

### Limitations

Our present work has two key limitations that inform the conclusions we are able to draw in the present manuscript and will guide our future work. First, our design relies on the assumption that the exposure is exogenous to the individual and not associated with pre-displacement health. This assumption could be violated if poor health increases the likelihood of being laid off during corporate downsizings^27^. This assumption is reasonable given the high rate of unemployment during the recession, but future work can test this assumption more thoroughly.

Secondly, we do not evaluate duration of unemployment or other labor market outcomes and instead focus only on the direct effect of the exogenous job loss shock. Previous work does indicate that labor market outcomes differ for displaced and non-displaced individuals for over a decade following displacement^10^. It is possible that labor market outcomes after displacement may be contributing to the excess mortality, but we do not evaluate this here. Instead, this is a direction for future work as we have elected to focus on the exogenous job loss to address endogeneity problems and draw causal conclusions on the effect of job loss; additional work could evaluate longer durations of unemployment as a moderator of the risk for mortality following job loss.

### Future Directions

Several studies investigated health, health behaviors, morbidity, and hospitalizations after job displacement. Displacement leads to increased risk of hospitalization due to traffic accidents, self-harm, and alcohol-related conditions, but not severe cardiovascular disease in 12 years of follow-up^28^. Displaced individuals are more likely to be depressed^29^, rate themselves as having poorer general health^29^, attempt suicide^15^, receive mental health or substance-use treatment^15^, have chronic physical health conditions^29^, and smoke cigarettes^26^. These varied results across causes of death and possible health effects are informative but broadly are hampered by small sample size, low precision, limited outcomes, and short follow up. Given the existing literature and the cause of death results identified in this analysis, it is necessary to evaluate morbidity and mechanisms leading to mortality in future work and disentangle the relationships between job loss, demographic factors, lifestyle factors and behaviors, and health. We can investigate disease incidence, survival after disease diagnosis, medication purchases, sickness absences, some lifestyle factors, and hospitalizations after displacement to understand the pathways from job displacement to cause-specific mortality.

### Conclusions

We have replicated and extended earlier research that suggests becoming unemployed is associated with all-cause and specific-cause mortality risk, up to 30 years after the job displacement. Our natural experiment design leverages exogenous job losses, where we investigated a sample of individuals who lost their job due to plant closure or mass layoff during the 1990s Finnish Recession and their similarly-employed peers who did not experience job displacement. We were able to control for a number of unobserved confounding factors with this design and demonstrate evidence consistent with a causal relationship between job loss and all-cause/specific-cause mortality risk. Future work can strengthen these causal conclusions by addressing health selection and genetic confounders, as well as follow up on significant specific causes of death with studies of lifestyle factors and health trajectories after displacement and prior to death.

## Supporting information

STROBE checklist

Supplemental Methods and Results

## Author Notes and Disclosures

### Data Sharing Statement

The research data used in this study are confidential and are not publicly available to protect participant privacy. Statistics Finland may release or grant permission to use confidential register data for scientific research, see the following website for more information: https://www.stat.fi/tup/tutkijapalvelut/index_en.html. For additional documentation or analysis code, contact the corresponding author (SZ).

### Ethics

The research ethical board of the Finnish Institute for Health and Welfare gave ethical approval for this work.

### Contributors

SZ data curation, investigation, formal analysis, methodology, project administration, visualization, original draft, review and editing, EA original draft, review and editing, AL conceptualization, resources, supervision, review and editing, JK conceptualization, resources, funding acquisition, supervision, review and editing, TM methodology, investigation, review and editing. SZ and TM have directly accessed the underlying data.

### Other Declarations

No medical writers nor editors contributed to this work. We did not use generative AI nor AI-assisted writing tools at any point in writing analysis code or writing the manuscript.

