## Supplementary material for "Causally-informative analyses of the effect of job displacement on all-cause and specific-cause mortality from the 1990s Finnish recession until 2020: A population registry study": STROBE checklist

STROBE Statement—checklist of items that should be included in reports of observational studies

|  | Item No. | Recommendation | Page  No. | Relevant text from manuscript |
| --- | --- | --- | --- | --- |
| **Title and abstract** | 1 | (*a*) Indicate the study’s design with a commonly used term in the title or the abstract | 1 | “job displacement” |
|  |  | (*b*) Provide in the abstract an informative and balanced summary of what was done and what was found | 2 |  |
| Introduction | | | |  |
| Background/rationale | 2 | Explain the scientific background and rationale for the investigation being reported | 4 |  |
| Objectives | 3 | State specific objectives, including any prespecified hypotheses | 5-6 | Here we capitalize on administrative data before, during, and after the 1990s recession in Finland to create a large cohort of involuntarily displaced individuals and non-displaced controls with 30 years of follow-up. Using this natural experiment, we evaluate the influence of job loss during the 1990s Finnish economic recession on mortality until 2020. Our work expands on prior research through the long duration of follow-up, which is crucial for diseases that may take longer to develop, as well as thorough investigation of specific causes of death. Lastly, we are able to utilize the entire Finnish population, as opposed to earlier works that needed to use a subsample of all Finnish residents14,16,17. Our research questions were as follows: 1) What is the relationship between job displacement during the 1990s Finnish economic recession and all-cause mortality until 2020? 2) What are the relationships between job displacement and specific causes of death? These analyses were pre-registered and the analysis plan is available at https://osf.io/bxydp/?view_only=63ce97108b5245eaafce2f083f59dec5. |
| Methods | | | |  |
| Study design | 4 | Present key elements of study design early in the paper | 6 | Here we capitalize on administrative data before, during, and after the 1990s recession in Finland to create a large cohort of involuntarily displaced individuals and non-displaced controls with 30 years of follow-up. Using this natural experiment, we evaluate the influence of job loss during the 1990s Finnish economic recession on mortality until 2020. |
| Setting | 5 | Describe the setting, locations, and relevant dates, including periods of recruitment, exposure, follow-up, and data collection |  |  |
| Participants | 6 | (*a*) *Cohort study*—Give the eligibility criteria, and the sources and methods of selection of participants. Describe methods of follow-up  *Case-control study*—Give the eligibility criteria, and the sources and methods of case ascertainment and control selection. Give the rationale for the choice of cases and controls  *Cross-sectional study*—Give the eligibility criteria, and the sources and methods of selection of participants | 6-8 |  |
|  |  | (*b*) *Cohort study*—For matched studies, give matching criteria and number of exposed and unexposed  *Case-control study*—For matched studies, give matching criteria and the number of controls per case | 10, Supplement for propensity score matching |  |
| Variables | 7 | Clearly define all outcomes, exposures, predictors, potential confounders, and effect modifiers. Give diagnostic criteria, if applicable | 6-10 |  |
| Data sources/ measurement | 8* | For each variable of interest, give sources of data and details of methods of assessment (measurement). Describe comparability of assessment methods if there is more than one group | 6-10 |  |
| Bias | 9 | Describe any efforts to address potential sources of bias | 7-8 | To define the exposure to be as free of confounds as possible, we operationalized job displacement as a binary variable indicating exogenous and involuntary job loss using information from linked employer-employee data. We define displaced workers as individuals separated from their private-sector jobs following plant closures or mass lay-offs of 50% or more employees. However, there are potential endogeneity issues due to firm restructuring. Therefore, a plant closure is not classified as genuine if a worker obtains a position within the same firm subsequently, or a significant proportion (70%) of displaced workers from the same plant relocate to another plant owned by the same firm within the next year^18^. This distinction enables us to differentiate between genuine plant closures and other organizational changes. Although job displacements typically do not accumulate for the same individuals, it is observed that less-skilled workers tend to match with weaker firms (e.g., Verho, 2020) and approximately 3% of displaced individuals in the sample experienced more than one displacement. To account for this potential bias, we focus only on the first displacement event for a given individual. This exposure definition has been validated in previous economic papers. |
| Study size | 10 | Explain how the study size was arrived at | 6 | We then utilized linked employee-employer data to restrict the sample to full-year (12 months of employment), non-student wage earners between the ages of 25-55 who had been employed continuously for at least two years in private-sector companies with at least 10 employees. |

| Quantitative variables | 11 | Explain how quantitative variables were handled in the analyses. If applicable, describe which groupings were chosen and why | 7-10 |
| --- | --- | --- | --- |
| Statistical methods | 12 | (*a*) Describe all statistical methods, including those used to control for confounding | 9-10, supplement |
|  |  | (*b*) Describe any methods used to examine subgroups and interactions | 9-10, supplement |
|  |  | (*c*) Explain how missing data were addressed | 9-10, supplement |
|  |  | (*d*) *Cohort study*—If applicable, explain how loss to follow-up was addressed  *Case-control study*—If applicable, explain how matching of cases and controls was addressed  *Cross-sectional study*—If applicable, describe analytical methods taking account of sampling strategy | 9-10, supplement |
|  |  | (*e*) Describe any sensitivity analyses | 10, supplement |
| Participants | 13* | (a) Report numbers of individuals at each stage of study—eg numbers potentially eligible, examined for eligibility, confirmed eligible, included in the study, completing follow-up, and analysed | Results |
|  |  | (b) Give reasons for non-participation at each stage | 10 |
|  |  | (c) Consider use of a flow diagram | 10 |
| Descriptive data | 14* | (a) Give characteristics of study participants (eg demographic, clinical, social) and information on exposures and potential confounders | N/A |
|  |  | (b) Indicate number of participants with missing data for each variable of interest | 10 |
|  |  | (c) *Cohort study*—Summarise follow-up time (eg, average and total amount) | 10 |
| Outcome data | 15* | *Cohort study*—Report numbers of outcome events or summary measures over time | 10-11 |
|  |  | *Case-control study—*Report numbers in each exposure category, or summary measures of exposure | 10-11 |
|  |  | *Cross-sectional study—*Report numbers of outcome events or summary measures | N/A |
| Main results | 16 | (*a*) Give unadjusted estimates and, if applicable, confounder-adjusted estimates and their precision (eg, 95% confidence interval). Make clear which confounders were adjusted for and why they were included | Table 2, 11, 8-9 |
|  |  | (*b*) Report category boundaries when continuous variables were categorized | 8-10, Supplement |
|  |  | (*c*) If relevant, consider translating estimates of relative risk into absolute risk for a meaningful time period | N/A |
|  |  |  | N/A |

Continued on next page

| Other analyses | 17 | Report other analyses done—eg analyses of subgroups and interactions, and sensitivity analyses | 11-13, Supplement |
| --- | --- | --- | --- |
| Discussion | | | |
| Key results | 18 | Summarise key results with reference to study objectives | 13 |
| Limitations | 19 | Discuss limitations of the study, taking into account sources of potential bias or imprecision. Discuss both direction and magnitude of any potential bias | 15 |
| Interpretation | 20 | Give a cautious overall interpretation of results considering objectives, limitations, multiplicity of analyses, results from similar studies, and other relevant evidence | 13-17 |
| Generalisability | 21 | Discuss the generalisability (external validity) of the study results | 13-17 |
| Other information | |  | |
| Funding | 22 | Give the source of funding and the role of the funders for the present study and, if applicable, for the original study on which the present article is based | 18 |

*Give information separately for cases and controls in case-control studies and, if applicable, for exposed and unexposed groups in cohort and cross-sectional studies.

**Note:** An Explanation and Elaboration article discusses each checklist item and gives methodological background and published examples of transparent reporting. The STROBE checklist is best used in conjunction with this article (freely available on the Web sites of PLoS Medicine at http://www.plosmedicine.org/, Annals of Internal Medicine at http://www.annals.org/, and Epidemiology at http://www.epidem.com/). Information on the STROBE Initiative is available at www.strobe-statement.org.
