## Supplemental Methods and Results for "Causally-informative analyses of the effect of job displacement on all-cause and specific-cause mortality from the 1990s Finnish recession until 2020: A population registry study"

*Measures*

Underlying cause of death in the Finnish Death Register is defined as “the disease which has initiated the series of illnesses leading directly to death, or the circumstances connected with an accident or an act of violence which caused the injury or poisoning leading to death”^1^. The Finnish Death Register used an adapted version of ICD-9 for deaths between 1987-1995, and ICD-10 for deaths occurring from 1996 onwards^2^. To facilitate longitudinal analyses, Statistics Finland has created a system of 54 main classes of diagnoses, which harmonizes causes of death across all classification systems utilized in the Death Register over time^2^. Table S1 provides a list of the 54 main classes and their corresponding ICD-10 codes, as well as how we grouped these classes for analysis.

Education was originally reported from the Statistics FOLK Basic data as an 8-category variable representing the educational level of the highest qualification or degree. This was then recoded into three values representing primary (original value of 1), secondary (original values 3 or 4), and tertiary education (original values 5, 6, 7, 8).

*Analyses*

To further strengthen causal inference, we utilized propensity score matching to create a set of matched controls for the treatment sample. We used the R package MatchIt^3^ to generate propensity scores and match controls to the treatment sample via nearest neighbor with a 4:1 ratio, without replacement^4^. To create the propensity scores, we utilized covariates that predicted both mortality and displacement: birth year, sex, year of assessment, and education. The 4:1 matching resulted in a total sample with N=571,285 individuals (N=114,257 treatment, N=457,028 controls). Table S3 shows a detailed view of the balance check and corresponds to Table 1 in the main text, showing the full sample.

We also conducted various sensitivity analyses stratifying on covariates of interest to evaluate whether the effect of displacement on all-cause mortality differed in some subgroup as compared to the full sample analysis. We ran stratified analyses by birth year, sex, education, and year of displacement.

Lastly, while we have date of death for the entire sample, we only have cause of death data on a subset of the deceased individuals, based on data permission for a larger project. Therefore, we compared demographic descriptives between deceased individuals with and without cause of death information. We also evaluated the effect of displacement on all-cause mortality in the full sample as compared to the sample of individuals with cause of death data, to ensure the effect of interest did not differ.

**Supplemental Results**

The full results of the all-cause mortality survival models in the propensity score matched sample are presented in Table S4. Overall, the results from the propensity score analysis agree with respect to effect significance and direction as compared to the analysis in the full sample. The effect sizes for job displacement are somewhat attenuated in the propensity-score matched sample as compared to the full sample, but the effect is significant at all durations of follow-up. The effects of covariates (sex, education, marital status, and having children under the age of 18) on mortality do not differ in magnitude in the propensity-score matched sample as compared to the full sample.

We ran stratified all-cause mortality models by birth year, sex, education, and year of displacement and compared those results to those from the full sample to further understand how covariates may impact the effect of job displacement on mortality. Full results are presented in Table S5 and are described in the main text as they compare to the non-stratified model.

Descriptives for the sample of individuals with cause of death data, as compared to the full sample, are presented in Table S6. Cause of death data was available for approximately 2/3rds of the deceased individuals. In general, individuals with available cause of death data had later birth years, had a younger mean age at death, were more likely to be male, and were slightly more likely to be displaced as compared to the sample without available cause of death data.

Table S7 presents the results of the all-cause morality model in the subset of individuals with cause of death information. The relationships between education-mortality and marital status-mortality did not change with respect to effect magnitude or significance as compared to the full sample. Hazard ratios for sex were significantly smaller (i.e., the effect of being a woman is more protective against mortality) in the subset analysis as compared to the full sample. On the other hand, having children under the age of 18 significantly predicted increased risk of all-cause mortality at all durations of follow-up in the subset analysis, whereas in the full sample analysis this was only the case for the earliest follow-up. Lastly, the effect of job displacement on mortality was comparable in terms of magnitude in the subset analysis as compared to the full sample. We therefore are confident that our specific cause of death results are generalizable, despite being from a subset of the full sample.

**Supplemental Tables**

| Table S1: Causes and Number of Deaths in Analyses | | | | |
| --- | --- | --- | --- | --- |
| Cause of Death as Evaluated in Analyses | **ICD-10**  **Code(s)** | **Statistics Finland Class** | **Total Deaths**  **by December**  **31^st^, 2020** | **% Deaths in Displaced Individuals** |
| All Cause | A00-Y89, U071, U072 | 1-54 | 93,199 (62,107 with specific cause data available) | 20.0% |
| Any Disease | A00-R99, X45, U071, U072 | 1-41 | 54,778 | 21.0% |
| Cancer | C00-D48 | 4-22 | 22,432 | 20.9% |
| Ischaemic Heart Disease | I20-I25 | 27 | 9,658 | 21.5% |
| Infectious and Respiratory Disease | A00-B99, J00-J99 | 1-3, 31-35 | 2,343 | 22.2% |
| Endocrine | E00-E90 | 23, 24 | 928 | 20.6% |
| Dementia and Nervous System | F01, F03, G30, R54, G00-G29, G31.0-G311, G31.8-G620 (pl. G4051), G622-G720, G722-H95 | 25, 26 | 3,329 | 18.4% |
| Any External Cause | V01-X44, X46-Y89 | 42-53 | 7,239 | 22.1% |
| Suicide | X60-X84, Y870 | 50 | 2,719 | 21.9% |
| Alcohol | F10, G312, G4051, G621, G721, I426, K292, K70, K860, K852, 0354, P043, Q860, X45 | 41 | 6,218 | 23.1% |
| Accidents | V01-X44, X46-X59, Y85-Y86, Y10-Y15 | 42-49 | 4,008 | 22.1% |
| Violence | X85-Y09, Y871 | 51 | 208 | 25.5% |
| No Death Certificate | R999 | 54 | 84 | 27.4% |
| Data Unavailable | - | - | 31,182 | 24.1% |

| Table S2: Effect of Job Displacement on Specific Cause Mortality | | | | |
| --- | --- | --- | --- | --- |
| HR [95% CI] | **Follow Up Duration** | | | |
|  | **1990-2000** | **1990-2005** | **1990-2010** | **1990-2020** |
| Any Disease | 1.15  [1.07, 1.22] | 1.14  [1.09, 1.19] | 1.13  [1.10, 1.15] | 1.09  [1.07, 1.11] |
| Cancer | 1.04  [0.94, 1.15] | 1.07 [0.99, 1.15] | 1.08  [1.03, 1.14] | 1.08  [1.05, 1.12] |
| Ischaemic Heart Disease | 1.04  [0.90, 1.21] | 1.11  [1.01, 1.23] | 1.11  [1.03, 1.20] | 1.08  [1.03, 1.14] |
| Infectious or Respiratory Disorder | 1.29  [0.90, 1.83] | 1.34  [1.06, 1.69] | 1.26  [1.14, 1.39] | 1.22  [1.11, 1.35] |
| Endocrine Disorder | 1.23  [0.74, 2.05] | 1.08  [0.76, 1.53] | 1.03  [0.79, 1.34] | 1.06  [0.91, 1.25] |
| Dementia or Nervous System Disease | 1.24  [0.79, 1.94] | 1.26  [0.96, 1.67] | 1.13  [0.93, 1.37] | 0.99  [0.91, 1.08] |
| Any External Cause | 1.39  [1.26, 1.55] | 1.26  [1.16, 1.37] | 1.25  [1.16, 1.34] | 1.19  [1.13, 1.26] |
| Suicide | 1.38  [1.19, 1.60] | 1.23  [1.09, 1.39] | 1.24  [1.16, 1.34] | 1.18  [1.08, 1.30] |
| Alcohol | 1.73  [1.48, 2.01] | 1.41  [1.26, 1.57] | 1.33  [1.22, 1.44] | 1.23  [1.16, 1.31] |
| Accidents | 1.46  [1.24, 1.70] | 1.27  [1.13, 1.43] | 1.26  [1.14, 1.39] | 1.19  [1.10, 1.28] |
| Violence | 1.76  [1.02, 3.04] | 1.91  [1.30, 2.83] | 1.70  [1.19, 2.44] | 1.68  [1.23, 2.30] |

| Table S3: Descriptives of Sample Across Possible Years of Exposure for Propensity Matched Sample | | | | | | | | | |  |
| --- | --- | --- | --- | --- | --- | --- | --- | --- | --- | --- |
|  | **1990** | | **1991** | | **1992** | | **1993** | | **1994** | |
|  | Matched Controls | Displaced | Matched Controls | Displaced | Matched Controls | Displaced | Matched Controls | Displaced | Matched Controls | Displaced |
| N  Individuals | 60,284 | 15,071 | 124,776 | 31,194 | 126,276 | 31,569 | 88,528 | 22,132 | 57,164 | 14,291 |
| %  Female | 43.4 | 43.3 | 37.8 | 37.8 | 37.0 | 37.0 | 39.3 | 39.3 | 43.5 | 43.5 |
| Age Mean (SD) | 39.2  (8.4) | 39.2  (8.4) | 39.3  (8.3) | 39.3  (8.3) | 39.4  (8.3) | 39.4  (8.3) | 40.0  (8.3) | 40.0  (8.3) | 40.4  (8.2) | 40.4  (8.2) |
| % Secondary Education Year N-1 | 41.9 | 41.9 | 42.8 | 42.8 | 40.4 | 40.4 | 38.9 | 38.9 | 37.1 | 37.1 |
| % Tertiary Education Year N-1 | 19.5 | 19.5 | 20.2 | 20.2 | 25.6 | 25.6 | 28.9 | 28.9 | 29.8 | 29.8 |
| % Married  Year N-1 | 63.8 | 62.1 | 63.0 | 60.5 | 62.4 | 61.5 | 62.4 | 61.4 | 62.0 | 61.2 |
| % Children Under 18 Year N-1 | 50.1 | 49.8 | 50.6 | 49.2 | 50.7 | 51.2 | 50.4 | 50.9 | 50.4 | 51.0 |

| Table S4: Results from All-Cause Mortality Survival Models in the Propensity Score Matched Sample | | | | |
| --- | --- | --- | --- | --- |
| HR [95% CI] | **Follow Up Duration** | | | |
|  | **1990-2000** | **1990-2005** | **1990-2010** | **1990-2020** |
| Displacement | 1.06  [1.00, 1.11] | 1.08  [1.04 1.12] | 1.08  [1.05, 1.12] | 1.07  [1.04, 1.09] |
| Sex | 0.41  [0.39, 0.44] | 0.42  [0.40, 0.44] | 0.43  [0.42, 0.45] | 0.47  [0.46, 0.48] |
| Secondary Education | 1.08  [1.01, 1.15] | 1.02  [0.97, 1.07] | 0.94  [0.91, 0.98] | 0.90  [0.87, 0.92] |
| Tertiary Education | 0.73  [0.68, 0.80] | 0.71  [0.67, 0.76] | 0.67  [0.64, 0.71] | 0.66  [0.64, 0.68] |
| Marital Status | 0.49  [0.46, 0.52] | 0.55  [0.52, 0.57] | 0.58  [0.56, 0.60] | 0.66  [0.64, 0.67] |
| Children Under 18 | 1.17  [1.10, 1.24] | 1.03  [0.99, 1.09] | 0.96  [0.93, 0.99] | 0.91  [0.89, 0.93] |

Note: The reference category for displacement was non-displaced individuals, the reference category for sex was male, the reference category for education was primary, the reference category for marital status was unmarried, and the reference category for children under 18 was no.

| Table S5: Effect of Job Displacement on Mortality from Stratified All-Cause Mortality Models | | | | | |
| --- | --- | --- | --- | --- | --- |
| Stratification Basis | **Sample** | **Follow Up Duration** | | | |
|  |  | **1990-2000** | **1990-2005** | **1990-2010** | **1990-2020** |
| None | Full  Sample | 1.17  [1.12, 1.23] | 1.15  [1.11, 1.18] | 1.12  [1.10, 1.15] | 1.09  [1.07, 1.10] |
| Sex | Males | 1.20  [1.14, 1.27] | 1.15  [1.11, 1.17] | 1.14  [1.10, 1.17] | 1.10  [1.08, 1.12] |
|  | Females | 1.08  [0.99, 1.19] | 1.12  [1.05, 1.19] | 1.09  [1.04, 1.14] | 1.05  [1.02, 1.08] |
| Education | Primary | 1.14  [1.07, 1.21] | 1.13  [1.08, 1.19] | 1.11  [1.07, 1.15] | 1.08  [1.05, 1.10] |
|  | Secondary | 1.20  [1.11, 1.30] | 1.16  [1.09, 1.22] | 1.14  [1.10, 1.19] | 1.09  [1.06, 1.12] |
|  | Tertiary | 1.23  [1.11, 1.39] | 1.16  [1.07, 1.26] | 1.13  [1.06, 1.20] | 1.10  [1.06, 1.14] |
| Birth Year | 1935-1946 | 1.07  [1.00, 1.14] | 1.07  [1.02, 1.12] | 1.05  [1.02, 1.09] | 1.05  [1.03, 1.08] |
|  | 1947-1958 | 1.28  [1.19, 1.38] | 1.21  [1.15, 1.28] | 1.19  [1.15, 1.24] | 1.12  [1.09, 1.15] |
|  | 1959-1969 | 1.27  [1.09, 1.46] | 1.23  [1.11, 1.37] | 1.21  [1.12, 1.31] | 1.12  [1.06, 1.18] |
| Displacement Year | 1990 | 1.19  [1.08, 1.31] | 1.19  [1.11, 1.27] | 1.14  [1.08, 1.20] | 1.08  [1.04, 1.13] |
|  | 1991 | 1.23  [1.15, 1.33] | 1.20  [1.13, 1.26] | 1.19  [1.14, 1.24] | 1.13  [1.10, 1.16] |
|  | 1992 | 1.13  [1.04, 1.23] | 1.12  [1.06, 1.19] | 1.09  [1.04, 1.14] | 1.06  [1.03, 1.10] |
|  | 1993 | 1.14  [1.02, 1.27] | 1.10  [1.02, 1.19] | 1.10  [1.04, 1.16] | 1.07  [1.04, 1.11] |
|  | 1994 | 1.05  [0.89, 1.23] | 1.00  [0.90, 1.12] | 1.01  [0.94, 1.10] | 1.05  [1.00, 1.10] |

| Table S6: Descriptive Comparison for Sample With and Without Cause of Death Data | | |
| --- | --- | --- |
|  | Cause of Death Data Unavailable | Cause of Death Data Available |
| N | 31,182 | 62,017 |
| % Female | 42.5% | 25.8% |
| % Displaced | 24.1% | 27.4% |
| Median Birth Year | 1940 | 1948 |
| Median Age at Death | 71 | 63 |

| Table S7: Results from All-Cause Mortality Survival Models in the Subset of Individuals with Cause of Death Information | | | | |
| --- | --- | --- | --- | --- |
| HR [95% CI] | **Follow Up Duration** | | | |
|  | **1990-2000** | **1990-2005** | **1990-2010** | **1990-2020** |
| Displacement | 1.21  [1.14, 1.28] | 1.17  [1.12 1.21] | 1.15  [1.12, 1.19] | 1.10  [1.08, 1.12] |
| Sex | 0.34  [0.32, 0.37] | 0.35  [0.22, 0.36] | 0.35  [0.34, 0.37] | 0.38  [0.37, 0.39] |
| Secondary Education | 1.07  [1.01, 1.13] | 1.03  [0.98, 1.07] | 0.97  [0.94, 1.00] | 0.91  [0.89, 0.93] |
| Tertiary Education | 0.66  [0.62, 0.72] | 0.68  [0.64, 0.71] | 0.65  [0.62, 0.67] | 0.63  [0.62, 0.65] |
| Marital Status | 0.47  [0.44, 0.59] | 0.51  [0.49, 0.53] | 0.54  [0.53, 0.56] | 0.60  [0.59, 0.62] |
| Children Under 18 | 1.29  [1.22, 1.36] | 1.23  [1.18, 1.27] | 1.18  [1.15, 1.22] | 1.17  [1.15, 1.19] |

Note: The reference category for displacement was non-displaced individuals, the reference category for sex was male, the reference category for education was primary, the reference category for marital status was unmarried, and the reference category for children under 18 was no.
